# HDL Particle Concentration and Size Predict Incident Coronary Artery Disease Events in People with Type 1 Diabetes

**DOI:** 10.1101/2023.11.06.23298165

**Authors:** Tina Costacou, Tomas Vaisar, Rachel G. Miller, W. Sean Davidson, Jay W. Heinecke, Trevor J. Orchard, Karin E. Bornfeldt

## Abstract

**Background:** Cholesterol efflux capacity (CEC) negatively correlates with cardiovascular disease risk. Small HDL particles account almost quantitively for CEC, perhaps mediated through efflux of outer leaflet plasma membrane phospholipids by ABCA1. People with type 1 diabetes (T1D) are at increased risk of coronary artery disease (CAD) despite normal levels of HDL-cholesterol (HDL-C). We therefore tested the hypotheses that small HDL particles (HDL-P)—rather than HDL-C levels—predict incident CAD in T1D.

**Methods:** Incident CAD (CAD death, myocardial infarction, and/or coronary revascularization) was determined in a cohort of 550 participants with childhood-onset T1D. HDL-P was quantified by calibrated ion mobility analysis. CEC and phospholipid efflux were quantified with validated assays.

**Results:** During a median follow-up of 26 years, 36.5% of the participants developed incident CAD. In multivariable Cox models, levels of HDL-C and apolipoprotein A-I (APOA1) did not predict CAD risk. In contrast, extra-small HDL particle levels strongly and negatively predicted risk (hazard ratio [HR]=0.25, 95% confidence interval [CI]=0.13–0.49). An increased concentration of total HDL particles (T-HDL-P) (HR=0.87, CI=0.82–0.92) and three other HDL sizes were weaker predictors of risk: small HDL (HR=0.80, 0.65-0.98), medium HDL (HR=0.78, CI=0.70–0.87) and large HDL (HR=0.72, CI=0.59–0.89). Although CEC negatively associated with incident CAD, that association disappeared after the model was adjusted for T-HDL-P. Isolated small HDLs strongly promoted ABCA1-dependent efflux of membrane outer leaflet phospholipids.

**Conclusions:** Low concentrations of T-HDL-P and all four sizes of HDL subpopulations predicted incident CAD independently of HDL-C, APOA1, and other common CVD risk factors. Extra-small HDL was a much stronger predictor of risk than the other HDLs. Our data are consistent with the proposal that small HDLs play a critical role in cardioprotection in T1D, which might be mediated by macrophage plasma membrane outer leaflet phospholipid export and cholesterol efflux by the ABCA1 pathway.

## Introduction

Low levels of HDL cholesterol (HDL-C) associate with increased risk of incident atherosclerotic cardiovascular disease (CVD) in the general population.^1, 2^ None the less, people with type 1 diabetes (T1D), who have normal or even elevated levels of HDL cholesterol (HDL-C), ^2,3^ also have an increased risk of atherosclerotic CVD.^3–5^ As a continuous variable, low levels of HDL-C associate with increased risk of incident CVD in the general population and in people with T1D.^1, 2^ Categorical analysis reveals a nonlinear association of HDL-C in T1D, with risk increasing in both the lowest (<50 mg/dL) and highest (>80 mg/dL) HDL-C groups.^2^ However, genetic variations that affect HDL-C levels do not associate with CVD in the general population,^6^ while randomized clinical trials of pharmacological interventions aimed at raising HDL-C have failed to reduce CVD in statin-treated patients.^7, 8^ Collectively, these observations indicate that HDL-C levels do not always predict CVD risk and that increasing HDL-C is not necessarily therapeutic. These discrepancies highlight a central problem in HDL biology: Is HDL cardioprotective or is HDL-C simply a marker for other risk factors?^9–11^ To address this issue, it is critical to identify HDL metrics that truly reflect CVD risk.

Multiple cardioprotective functions have been proposed for HDL, ranging from suppression of endothelial cell inflammation to regulation of complement activation.^11, 12^ The best validated model of HDL’s proposed cardioprotection is the ability to promote cholesterol efflux from cholesterol-laden macrophages in the artery wall.^9, 10^ The first step in this pathway involves ABCA1, an ATP-binding cassette transporter that is highly induced when macrophages accumulate cholesterol.^9, 10, 13^ Studies from large clinical populations support this model: cholesterol efflux capacity (CEC)—the ability of serum HDL (APOB-depleted serum) to promote cholesterol efflux from macrophages expressing ABCA1—predicts CVD in humans.^11, 14–16^ Importantly, the association of CEC with CVD risk is independent of HDL-C and levels of APOA1, the major structural protein of HDL. Taken together, these observations suggest that serum HDL’s capacity to promote cholesterol efflux from macrophages *in vitro* reflects its functionality *in vivo*.

Human HDL is a complex mixture of particles ranging from ∼7 nm to ∼14 nm in diameter.^17^ Because spherical HDL particles can vary over 6-fold in cholesterol content per particle, it is not surprising that HDL-C levels provide limited information about HDL’s cardioprotective effects. Two potentially more useful metrics are HDL particle number (HDL-P, the concentration of total HDL particles) and the concentrations of specific subpopulations of HDL as defined by size.^16^

Many lines of evidence indicate that small HDL particles promote CEC by the ABCA1 pathway more effectively than large HDL particles.^18, 19^ We used molecular modeling, reconstituted HDLs, and chemical crosslinking to demonstrate that the underlying mechanism involves the C-terminal ends of APOA1.^20^ In small HDLs, the C-termini of APOA1 are “flipped” off the particle’s lipid surface, increasing the engagement of APOA1 with ABCA1.^21^ In contrast, APOA1 of larger HDLs is unable to interact productively with ABCA1 because it forms a helical bundle that binds to lipid on the particle. Thus, extra-small and small HDLs may be key mediators and indicators of HDL’s cardioprotective effects mediated through ABCA1.

A widely accepted mechanism for promoting cellular cholesterol efflux by the ABCA1 pathway is the alternating access model, which flops phospholipids (PLs) from the inner leaflet to the outer leaflet of the plasma membrane.^22^ Phospholipid translocation drives cholesterol efflux from the plasma membrane.^22^ Based on molecular modeling, structural analysis, and studies of cells expressing wild-type and mutant ABCA1, we proposed a different mechanism: cholesterol efflux by ABCA1 involves the export of phospholipids from the outer leaflet of the membrane into the extracellular milieu.^21^ However, the relative roles of inner leaflet and outer leaflet PLs as substrates for export in cells expressing ABCA1 have yet to be determined.

In the current study, we tested the hypothesis that HDL particle size and concentration are key determinants of cardioprotection in people with T1D, a population at high risk for CVD despite the normal/high HDL-C concentrations. We quantified HDL-P by calibrated ion mobility analysis (IMA) and assessed ABCA1 CEC and macrophage CEC with validated assays. We also used cells with inducible ABCA1 to determine the relative roles of inner and outer leaflet PLs as substrates for export by ABCA1 to the different sizes of HDL.

## Methods

### Study population

The Re-Evaluating The Role of HDL Cholesterol in coronary artery disease (RETRO HDLc) study^23^ is an ancillary study of the Epidemiology of Diabetes Complications (EDC) study,^24, 25^ which prospectively followed participants with childhood-onset T1D (<17 years old) for the development of diabetic complications. EDC study participants were diagnosed with T1D (or seen within 1 year of diagnosis) at Children’s Hospital of Pittsburgh in 1950–1980. They were prospectively followed with biennial surveys and examinations (1986–1998). Subsequently, biennial surveys continued, and further examinations were performed after 18, 25, and 30 years of follow-up. The University of Pittsburgh Institutional Review Board approved all study protocols.

### Survey and examination data

Demographic, healthcare, diabetes self-care, and medical history information was self-reported. Participants who had smoked ≥100 cigarettes over their lifetime were classified as “ever smokers.” Anthropometrics and fasting blood and urine samples were obtained during the clinical examinations, and samples were stored at −70° C. Body mass index (BMI) was calculated. Blood pressure was measured with a random zero sphygmomanometer following a 5-minute rest,^26^ and hypertension was defined as blood pressure ≥140/90 mmHg or antihypertensive medication use.

CAD was defined as myocardial infarction confirmed by Q-waves on electrocardiogram (Minnesota codes 1.1 or 1.2) or hospital records, coronary revascularization, or CAD death. In addition to death certificates, we obtained medical records, autopsy/coroners’ reports, and/or interviews with next-of-kin regarding the circumstances surrounding the death when possible. The underlying cause of death was determined and classified as cardiovascular (subdivided into coronary artery, cerebrovascular, or other), renal, acute complication, accident/suicide, infection, cancer, other diabetes-related, or other non-diabetes-related by a physician-based Mortality Classification Committee following The Diabetes Epidemiology Research International procedures.^27^

### Laboratory measurements

Stable glycated hemoglobin (HbA_1_) was measured by ion exchange chromatography (Isolab, Akron, OH) for the first 18 months; thereafter, automated high-performance liquid chromatography (Diamat; Bio-Rad, Hercules, CA) was used. The two assays were highly correlated (r=0.95). The original HbA_1_ values were converted to DCCT-aligned HbA_1c_ values using regression formulae derived from duplicate analyses (DCCT HbA_1c_ = (0.83 * EDC HbA_1_) + 0.14). HDL-C was determined using ultracentrifugation precipitation techniques.^28^ Total cholesterol and triglycerides were measured enzymatically.^29, 30^ Non-HDL-C was calculated as total cholesterol minus HDL-C. Apolipoprotein A-I (APOA1) was determined via immunoelectrophoresis.^31^ Serum and urinary albumin,^32^ serum creatinine (Ectachem 400 Analyzer, Eastman Kodak Co., Rochester, NY), and white blood cell counts (WBC, S-plus IV counter, Coulter Electronics, Hialeah, FL) were also measured. Glomerular filtration rate was estimated (eGFR) by the CKD EPI equation.^33^

### Quantification of HDL particle concentration (HDL-P)

HDL-P and CEC were assessed using the first available stored plasma specimen obtained before incident CAD or at the end of follow-up for non-cases. For ∼95% of the participants, a sample was available from 1986–90. For 5%, only a sample from a later examination was available. HDL-P was quantified by calibrated IMA on a differential ion mobility analyzer (DMA, TSI Inc., MN). Total HDL particle concentration (T-HDL-P) and four major HDL subpopulations (extra-small [XS-HDL-P], 7.6 nm diameter; small [S-HDL], 8.2 nm diameter; medium [M-HDL], 9.1 nm diameter; and large [L-HDL], 10.9 nm diameter) were fitted to the IMA profiles by unsupervised, iterative curve fitting.^34^

### Quantification of CEC

HDL CEC was quantified with serum HDL (plasma-derived serum depleted of apolipoprotein B lipoproteins) using cells labeled with [^3^H]cholesterol.^14, 19, 35, 36^ All assays were carried out under conditions where CEC was a linear function of incubation time and serum HDL concentration. Macrophage CEC was quantified with J774 macrophages stimulated with cAMP and was calculated as the percentage of total [^3^H]cholesterol (medium plus cell-associated) released into the medium containing serum HDL minus that of cells incubated with medium alone. ABCA1 CEC was quantified using BHK (baby hamster kidney) cells with mifepristone-inducible human ABCA1 expression. ABCA1 CEC was calculated as the percentage of total [H^3^]cholesterol (medium plus cell-associated) released into the medium of BHK cells stimulated with mifepristone minus that of non-stimulated cells. Analyses were normalized to a pool of control HDL that was included in each batch of assays. Laboratory personnel were blinded to participants’ CAD status.

### Isolation of HDL subpopulations

HDL was isolated from 5 mL plasma obtained from a healthy volunteer who provided informed consent. The study was approved by the University of Washington Institutional Review Board (00012123). Plasma was quickly thawed at 37°C, and subjected to sequential ultracentrifugation to isolate HDL (density 1.063–1.210 g/mL) ^37^. The protein concentration in HDL was measured using the Bradford assay, with albumin as the standard. HDL (Total-HDL) was separated (0.3 mL/min) by sequential fractionation on three tandem Superdex 200 Increase 10/300 GL columns at 4°C into small-HDLs (raging from 7.7 to 9.0 nm), medium-HDLs (8.1-9.9 nm), and large-HDLs (9.8-11.9 nm).^38^ Each fraction was concentrated, and the molar concentrations and sizes of HDLs were determined by calibrated IMA.^19^

### Quantification of ABCA1-dependent export of outer leaflet phosphatidylcholine

Cyclodextrins are widely used to exchange membrane and medium lipids.^39^ Because the hydrophobic cavity of methyl-α-cyclodextrin (MαCD; AraChem, Tilburg, the Netherlands) does not bind cholesterol,^39^ we used it to selectively replace endogenous sphingolipids and phospholipids in the outer plasma membrane leaflets of BHK cells with 1-palmitoyl-2-oleoyl-sn-glycero-3-phosphocholine (POPC) and brain sphingomyelin (Avanti Polar Lipids, Inc.).^39^ Phosphatidylcholine (PC) was labeled by incubating cells with ^3^H-choline (^3^H-choline chloride, 77.5 Ci/mmol, PerkinElmer, NET109001MC) for 24 h. After washing the cells, we exchanged labeled PC in the outer leaflet of the plasma membrane with POPC (Avanti Polar Lipids, Inc.) by incubating the cells for 30 minutes with 40 mM MαCD, 1.5 mM POPC, and 1.5 mM brain sphingomyelin, as described by Li et al.^39^ The ratio of POPC to sphingomyelin used for lipid exchange was based on the reported concentration of PC and sphingomyelin in the plasma membrane of BHK cells^40^ and on pilot studies showing lack of detectable cell death under these conditions. ABCA1-mediated CEC was quantified using BHK cells, as described above for HDL CEC. Export of radiolabeled PC was quantified by thin layer chromatography separation of PLs in the media and scintillation counting.^39^

### Statistical analysis

Covariate data came from the examination in which HDL-P and CEC were measured. Pearson or Spearman correlation coefficients, as appropriate, were employed to evaluate factors related to these HDL metrics. Descriptive statistics were used to assess differences in participant characteristics by CAD incidence.

Separate Cox proportional hazards models with backward elimination were constructed for HDL-C and each novel HDL metric as the main independent variable, allowing for factors significantly associated with CAD univariately. Survival time was defined as the time in years from novel HDL metric and covariate assessment to the date of a first CAD event or, for non-cases, the last available follow-up. In addition to the univariate model, three multivariable models were constructed: the first allowed for lipid risk factors (non-HDL-C and HDL-C), the second allowed for lipid risk factors in addition to all other traditional CVD risk factors, and the third model allowed for variables in Model 2 and T-HDL-P (for models with HDL-C or CEC as the main independent variable) or CEC (for models with HDL-P as the main independent variable). Multivariable analyses were repeated, replacing non-HDL-C with LDL-C, triglycerides, and APOA1 in the smaller number of study participants who had fasted.

The proportional hazards assumption was assessed with an interaction term between each covariate of interest and logarithmically transformed time, keeping the fixed covariates in the model. For those covariates that violated the proportionality assumption, the covariate’s interaction with time (covariate * log(time)), in addition to its fixed term, was included in the models. Hazard ratios (HR) are presented per one SD increase for CEC, per 1 μmol/L increase for variables reflecting HDL-P and per 1 mg/dL increase for HDL-C. Two-sided p-values <0.05 were considered statistically significant. All analyses were conducted using SAS version 9.4 (SAS Institute, Cary, NC).

ABCA1-mediated lipid efflux experiments were performed in triplicate and analyzed by two-way ANOVA followed by Tukey’s multiple comparison tests.

## Results

### Characterization of study participants with and without CAD

Samples that could be analyzed for HDL-P and CEC were available from 85.4% of the 644 study participants who were free of CAD at study entry. Characteristics of those 550 people are listed in **Table 1**. During a median follow-up of 26 years, there were 201 (36.5%) incident CAD cases. At baseline, participants who subsequently developed CAD were older, had suffered from diabetes for a longer period, used a lower insulin dose per kg body weight, and were more likely to have hypertension. They also had higher white blood cell counts and adverse kidney disease profiles as well as higher non-HDL cholesterol and triglyceride levels. In contrast, the two groups had similar baseline levels of HDL-C and APOA1.

**Table 1.** Baseline characteristics of study participants with type 1 diabetes.

|  | **No CAD**<br>**n=349** | **Incident CAD**<br>**n=201** | **p-value** |
| --- | --- | --- | --- |
| Age (years) | 24.8 (20.8, 30.7) | 32.0 (26.5, 36.3) | **<0.0001** |
| Age at diabetes onset (years) | 8.2 (5.1, 11.7) | 8.7 (4.8, 11.2) | 0.98 |
| Diabetes duration (years) | 16.6 (11.7, 22.0) | 23.6 (18.2, 29.1) | **<0.0001** |
| Follow-up time (years) | 26.4 (16.2, 30.0) | 14.7 (8.0, 22.6) | **<0.0001** |
| Females (%; n) | 49.9 (174) | 48.3 (97) | 0.72 |
| Body mass index (kg/m<sup>2</sup>; n=347, 200) | 23.3 (21.1, 25.4) | 23.7 (21.9, 26.2) | 0.06 |
| Waist to hip ratio (n=329, 191) | 0.82 (0.76, 0.86) | 0.84 (0.79, 0.89) | **0.0004** |
| Ever smoker (%; n=346, 200) | 31.2 (108) | 47.5 (95) | **0.0001** |
| Insulin dose per body weight (n=330, 195) | 0.79 (0.63, 0.94) | 0.71 (0.58, 0.85) | **0.0007** |
| HbA<sub>1c</sub> (%) | 8.5 (7.7, 9.7) | 8.6 (7.8, 9.8) | 0.61 |
| Systolic blood pressure (mm Hg) | 108 (102, 116) | 114 (106, 124) | **<0.0001** |
| Diastolic blood pressure (mm Hg) | 70 (65, 78) | 74 (67, 82) | **0.0004** |
| Hypertension medications (%; n=334, 196) | 4.8 (16) | 16.8 (33) | **<0.0001** |
| Pulse (beats/min; n=349, 200) | 76 (70, 82) | 80 (72, 84) | **0.003** |
| Hypertension (%; n) | 9.7 (34) | 22.4 (45) | **<0.0001** |
| HDL cholesterol (mg/dL; n=349, 200) | 52.7 (45.8, 61.8) | 51.0 (44.9, 59.4) | 0.11 |
| Non-HDL cholesterol (mg/dL; n=349, 200) | 121.2 (101.5, 142.2) | 152.0 (124.4, 178.2) | **<0.0001** |
| Lipid medications (%; n=337, 196) | 0.0 (0) | 1.5 (3) | 0.05 |
| Triglycerides (mg/dL; n=335, 192) | 77.0 (55.0, 103.0) | 96.5 (72.5, 149.0) | **<0.0001** |
| Apolipoprotein A-I (mg/dL; n=343, 200) | 136.9 (19.5) | 140.2 (18.5) | 0.05 |
| ACE / ARB (%; n=337, 196) | 3.3 (11) | 5.6 (11) | 0.19 |
| White blood cell count (x10<sup>3</sup>/mm<sup>2</sup>; n=348, 201) | 5.9 (5.2, 7.3) | 6.8 (5.8, 8.2) | **<0.0001** |
| AER (μg/min; n=345, 201) | 10.8 (6.2, 51.9) | 36.8 (10.2, 476.7) | **<0.0001** |
| eGFR (mL/min/1.73 m<sup>2</sup>; n=349, 200) | 110.9 (91.4, 127.6) | 102.2 (78.4, 120.4) | **<0.0001** |
| Cholesterol efflux capacity (%) |  |  |  |
| Macrophages | 1.54 (0.43) | 1.51 (0.47) | 0.53 |
| ABCA1 | 1.34 (0.30) | 1.36 (0.33) | 0.43 |
| HDL-P concentration (μmol/L) |  |  |  |
| T-HDL-P | 7.29 (2.29) | 6.37 (2.67) | **<0.0001** |
| XS-HDL-P | 0.43 (0.30, 0.57) | 0.39 (0.25, 0.53) | **0.004** |
| S-HDL-P | 1.51 (1.07, 1.95) | 1.39 (0.84, 1.98) | 0.14 |
| M-HDL-P | 3.33 (2.61, 4.0) | 2.96 (2.09, 3.70) | **0.0003** |
| L-HDL-P | 1.23 (0.88, 1.92) | 0.99 (0.66, 1.56) | **<0.0001** |

Significant p-values (<0.05) are highlighted in bold. Data are percent (n), mean (SD), or median (25<sup>th</sup>, 75<sup>th</sup> percentile).
ACE, angiotensin converting enzyme inhibitor; AER, albumin excretion rate; ARB, angiotensin-receptor blocker; eGFR, estimated glomerular filtration rate; L-HDL-P, large HDL particle concentration; M-HDL-P, medium HDL particle concentration; S-HDL-P, small HDL particle concentration; T-HDL-P, total HDL particle concentration; XS-HDL-P, extra-small HDL particle concentration.

We next compared the two groups for baseline HDL particle concentration (HDL-P) and CEC. Total HDL-P (T-HDL-P) is the sum of the concentrations of four specific, size-defined, subpopulations of HDL in plasma, namely extra-small HDL (XS-HDL-P, ∼7.6 nm), small HDL (S-HDL-P, ∼8.2 nm), medium HDL (M-HDL-P, ∼9.1 nm), and large HDL (L-HDL-P, ∼10.9 nm). Univariately, T-HDL-P (p<0.0001) and the concentrations of XS-HDL-P, M-HDL-P, and L-HDL-P were all significantly (p≤0.004) lower in participants with incident CAD than in the non-cases (**Table 1**). S-HDL-P did not differ significantly among the groups (p=0.14). Moreover, HDL functionality—measured as macrophage CEC or ABCA1 CEC—did not differ by incident CAD status in univariate analysis (macrophage CEC, p=0.53; ABCA1 CEC, p=0.43). Supplemental Figure 1 depicts the distributions of HDL-C, HDL-P, and CEC by incident CAD status.

### Association of HDL metrics with incident CAD risk

Results from univariate and multivariable survival analysis are shown in **Table 2**. In the univariate models, the concentrations of T-HDL-P and three HDL subpopulations (XS-HDL-P, M-HDL-P, and L-HDL-P) demonstrated strong negative associations with incident CAD (HR 0.78-0.42). Allowing for non-HDL-C and HDL-C (Model 1) appeared to strengthen the associations, particularly for XS-HDL-P (HR=0.42 to HR=0.30). It also produced significant results for S-HDL-P. The negative associations of total, medium, and large HDL subpopulations remained significant. Moreover, associations of T-HDL-P and all the HDL-P subpopulations remained highly significant after allowing for all other traditional CAD risk factors that differed significantly between the CAD cases and non-cases (Model 2). This was especially true for XS-HDL, where we observed a 75% reduction in CAD risk per 1 μmol/L increase in the concentration of XS-HDL-P (HR=0.25, 95% CI 0.13–0.49) (**Figure 1**).

**Table 2.** Cox proportional hazards models^a^ (HR, 95% CI) for predicting incident CAD (n=537; 197 events)

| **Main independent variable** | **Univariate model** | **Model 1** | **Model 2** |
| --- | --- | --- | --- |
| HDL-C | **0.98 (0.97-0.995)** | 0.99 (0.98-1.00) | 0.99 (0.98-1.00) |
| T-HDL-P | **0.78 (0.72-0.86)** | **0.87 (0.82-0.92)** | **0.87 (0.82-0.92)** |
| XS-HDL-P | **0.42 (0.21-0.83)** | **0.30 (0.15-0.57)** | **0.25 (0.13-0.49)** |
| S-HDL-P | 0.83 (0.67-1.01) | **0.78 (0.64-0.95)** | **0.80 (0.65-0.98)** |
| M-HDL-P | **0.78 (0.70-0.87)** | **0.79 (0.71-0.89)** | **0.78 (0.70-0.87)** |
| L-HDL-P | **0.61 (0.49-0.75)** | **0.73 (0.59-0.90)** | **0.72 (0.59-0.89)** |
| Macrophage CEC <sup>b</sup> | 0.98 (0.85-1.13) | **0.76 (0.66-0.87)** | **0.83 (0.72-0.96)** |
| ABCA1 CEC <sup>b</sup> | 1.06 (0.92-1.22) | **0.83 (0.72-0.96)** | **0.82 (0.70-0.96)** |

<sup>a</sup> No adjustments were made in the univariate model.
Significant HRs and CIs are highlighted in bold.
**Model 1** allowed for HDL-C (for models where HDL-C was not the main independent variable) and non-HDL-C.
**Model 2** allowed for variables in Model 1 in addition to diabetes duration, sex, body mass index, smoking, HbA1c, hypertension (time-dependent variable and fixed term), white blood cell count, estimated glomerular filtration rate, and albumin excretion rate.
<sup>b</sup> HRs per SD (macrophage CEC, SD=0.44. ABCA1 CEC, SD=0.31)

**Figure 1.**
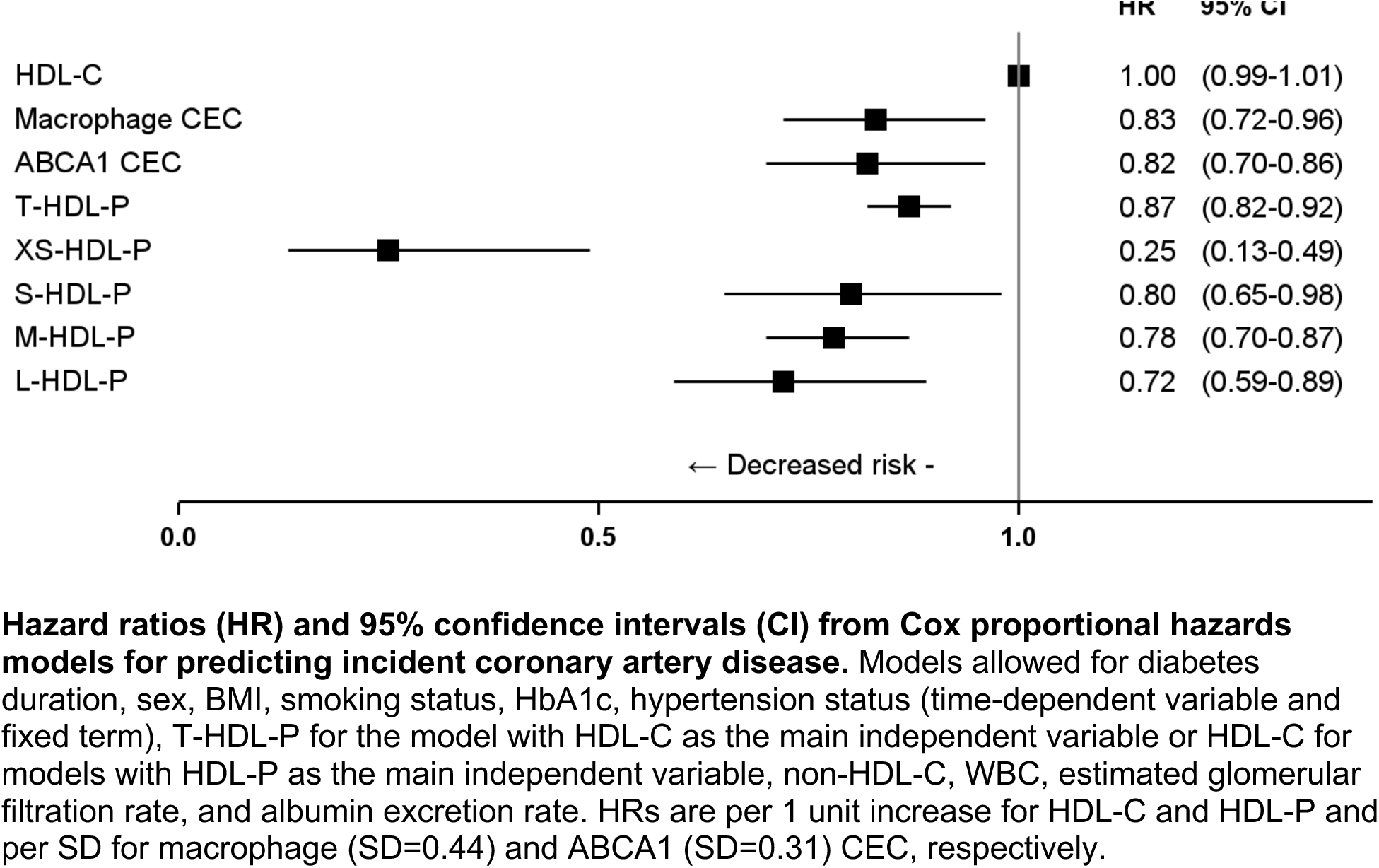
HDL particle concentration negatively predicts CAD risk in people with T1D.

In the univariate models, neither macrophage CEC nor ABCA1 CEC associated with CAD incidence. However, adjusting for non-HDL-C, HDL-C (Model 1), and other traditional CAD risk factors (Model 2) resulted in significant associations for both measures of serum HDL CEC (**Table 2**). However, after further adjustment for T-HDL-P (Model 3), both macrophage CEC and ABCA1 CEC lost significance (HR 0.97, CI 0.83-1.17 and HR 0.97, CI 0.81-1.15 respectively), strongly suggesting that CEC is not independent of HDL-P. Importantly, despite the strong association of higher levels of HDL-P, and especially XS-HDL, with lower incident CAD, HDL-C did not predict incident CAD (**Figure 1**). We obtained similar results when we replaced non-HDL-C with levels of LDL-C, triglycerides, and APOA1 in the fully adjusted model (Supplemental Table 1).

### Correlations between HDL metrics and other lipid risk factors for CAD

A key issue is whether HDL-P is independent of other lipid risk factors for CAD. We found that L-HDL-P correlated moderately with HDL-C (r=0.67, p<0.0001) while the correlation of T-HDL-P with HDL-C was weaker (r=0.43, p<0.0001), which is consistent with the much higher cholesterol content of large HDLs (**Table 3**). HDL-C significantly correlated with macrophage CEC (r=0.22, p<0.0001) but not with ABCA1 CEC (r=0.05, p=0.24), suggesting that pathways other than ABCA1 promote cholesterol efflux from macrophages. There was no correlation between HDL-C and XS-HDL-P (r=0.03, p=0.47).

**Table 3.** Correlation coefficients (p-values) among HDL-P, CEC and traditional lipid metrics.

|  | Cholesterol efflux capacity (%) |  | HDL-P concentration (μM/L) |  |  |  |  |
| --- | --- | --- | --- | --- | --- | --- | --- |
|  | Macrophage | ABCA1 | T-HDL-P | XS-HDL-P | S-HDL-P | M-HDL-P | L-HDL-P |
| HDL-C | 0.22 (**<0.0001**) | 0.05 (0.24) | 0.43<br>(**<0.0001**) | 0.03 (0.47) | -0.07 (0.11) | 0.27<br>(**<0.0001**) | 0.67 (**<0.0001**) |
| Non-HDL-C | 0.22 (**<0.0001**) | 0.27<br>(**<0.0001**) | -0.18<br>(**<0.0001**) | -0.06 (0.19) | 0.04 (0.35) | -0.12<br>(**0.002**) | -0.32 (**<0.0001**) |
| Triglycerides | 0.21 (**<0.0001**) | 0.21<br>(**<0.0001**) | -0.14<br>(**0.0009**) | -0.01 (0.82) | 0.14 (**0.001**) | -0.07 (0.09) | -0.38 (**<0.0001**) |
| APOA1 | 0.20 (**<0.0001**) | 0.08 (0.05) | 0.18<br>(**<0.0001**) | 0.06 (0.16) | 0.03 (0.55) | 0.11<br>(**0.008**) | 0.15<br>(**0.0003**) |
| Macrophage CEC | -- | 0.75<br>(**<0.0001**) | 0.35<br>(**<0.0001**) | 0.22<br>(**<0.0001**) | 0.37<br>(**<0.0001**) | 0.32<br>(**<0.0001**) | 0.17 (**<0.0001**) |
| ABCA1 CEC |  | -- | 0.30<br>(**<0.0001**) | 0.29<br>(**<0.0001**) | 0.38<br>(**<0.0001**) | 0.34<br>(**<0.0001**) | 0.07 (0.10) |
| T-HDL-P |  |  | -- | 0.46<br>(**<0.0001**) | 0.63<br>(**<0.0001**) | 0.88<br>(**<0.0001**) | 0.72 (**<0.0001**) |
| XS-HDL-P |  |  |  | -- | 0.41<br>(**<0.0001**) | 0.40<br>(**<0.0001**) | 0.23 (**<0.0001**) |
| S-HDL-P |  |  |  |  | -- | 0.54<br>(**<0.0001**) | 0.12 (**0.004**) |
| M-HDL-P |  |  |  |  |  | -- | 0.52 (**<0.0001**) |

Significant correlations (p<0.05) are highlighted in bold. ABCA1, ABCA1-specific serum HDL cholesterol efflux capacity (CEC); AER, albumin excretion rate; eGFR, estimated glomerular filtration rate; J774, J774 macrophage serum HDL CEC; L-HDL-P, large HDL particle concentration; M-HDL-P, medium HDL particle.

Macrophage CEC and ABCA1 CEC were strongly correlated (r=0.75, p<0.0001), which is consistent with the proposal that ABCA1 is the major pathway for HDL-dependent cholesterol efflux from macrophages. Macrophage CEC and ABCA1 CEC also correlated significantly with the concentration of each HDL subpopulation (macrophage CEC, r=0.17-0.37; ABCA1 CEC, r=0.29-0.38, all p-values<0.0001) except for L-HDL-P, which was not significantly correlated with ABCA1 CEC (r=0.07, p=0.10). This is consistent with the finding that L-HDL was less able to induce cholesterol efflux than smaller HDLs.^18, 19^ Macrophage CEC, but not ABCA1 CEC (r=0.22, p<0.0001 and r=0.05, p=0.24, respectively), weakly correlated with HDL-C, likely because cholesterol efflux from macrophages involves two mechanisms distinct from the ABCA1 pathway: the ABCG1 pathway, and diffusion.^14^

Non-HDL-C also correlated weakly with macrophage CEC (r=0.22, p<0.0001) and ABCA1 CEC (r=0.27, p<0.0001). Non-HDL-C correlated moderately and negatively with L-HDL-P (r=−0.32, p<0.0001) and weakly with M-HDL (r=−0.12, p=0.002), but was not correlated with XS-HDL-P and S-HDL-P. APOA1 levels correlated weakly with macrophage CEC and T-HDL-P, M-HDL-P, and L-HDL-P (r ≤0.20, p≤0.008) for all variables. These data suggest that HDL particle concentration and size—particularly for XS-HDL-P—are independent of traditional CAD risk factors.

Correlations between other clinical characteristics and HDL metrics are shown in Supplemental Table 2. HbA1c levels correlated positively and modestly with macrophage CEC (r=0.22, p<0.0001) and ABCA1 CEC (r=0.18, p<0.0001). Albumin excretion rate correlated negatively with T-HDL-P (r=−0.23, p<0.0001), M-HDL-P (r=−0.23, p<0.0001), and L-HDL-P (r=−0.25, p<0.0001). Other correlations were also weak.

### Small HDLs selectively and potently promote phosphatidylcholine export from the outer leaflet of cells expressing ABCA1

HDL isolated by ultracentrifugation from freshly thawed plasma (stored at −80 °C after collection) was separated by high-resolution size exclusion chromatography and three fractions were collected: small-HDLs (raging from 7.7 to 9.0 nm), medium-HDLs (8.1-9.9 nm), and large-HDLs (9.8-11.9 nm). For each isolated fraction and for total HDL, we used calibrated ion mobility analysis to quantify HDL particle concentration and size (HDL-P)^34^. Representative size profiles of the HDL populations are shown in Figure S2. All functional analyses were performed at the same molar concentration of HDL particles (80 nM) in each assay.

PL export was quantified as radiolabeled PC in the medium after BHK cells expressing ABCA1 were incubated for 4 h with APOA1 or equimolar concentrations of the three sizes of isolated HDL (**Figure 2A**). APOA1 and small HDLs potently stimulated PC export from the cells. Small HDLs were about 50% as active as lipid-free APOA1 in promoting PC efflux. In contrast, medium HDLs were less than 10% as effective as APOA1 in promoting PC export, and large HDLs failed to promote PC export.

**Figure 2.**
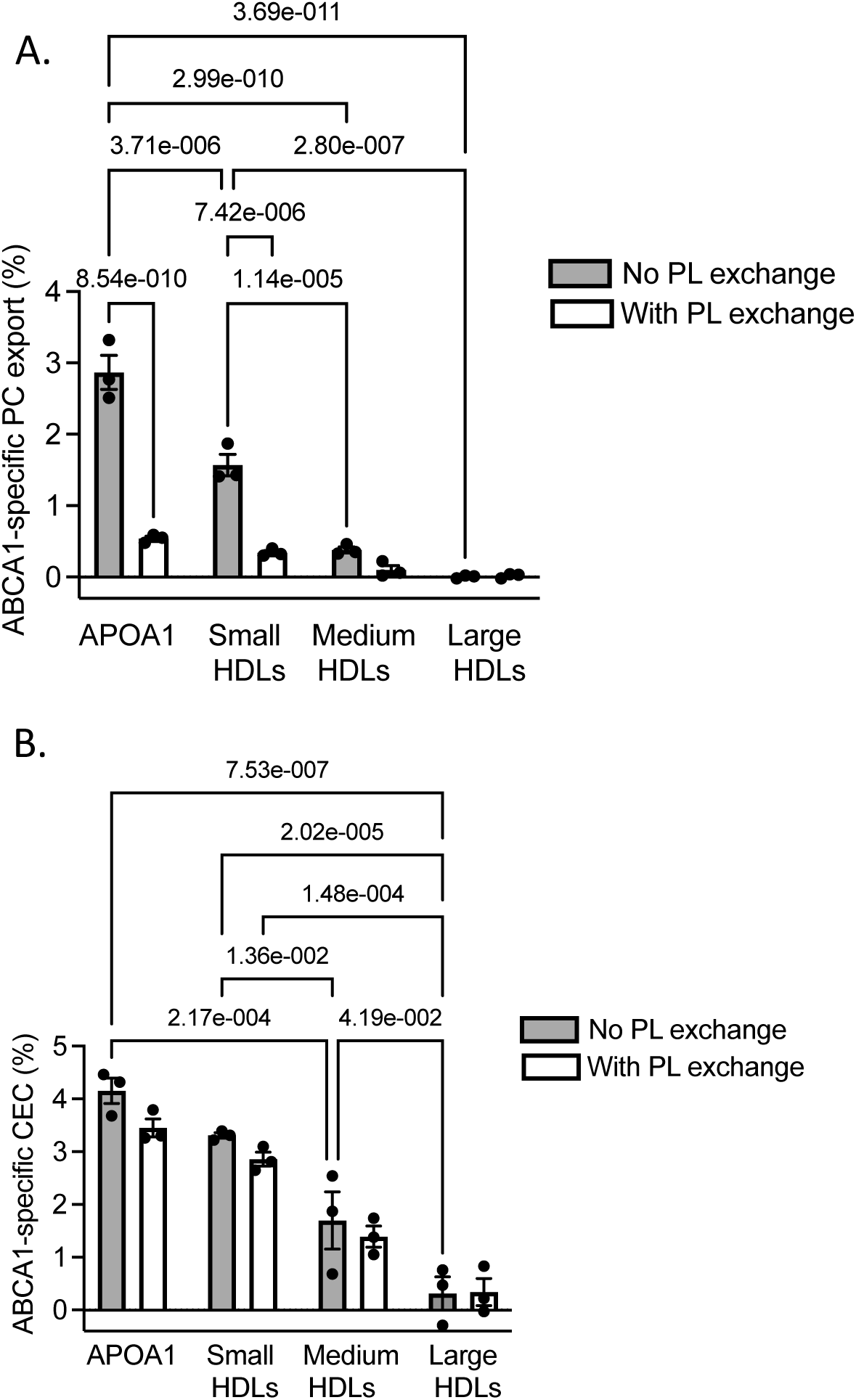
ABCA1-dependent export of membrane outer leaflet PCs by different sizes of HDL. (**A**) Phosphatidylcholine (PC) export and (**B**) cholesterol efflux capacity (CEC) by ABCA1 expressing cells were quantified using equimolar concentrations (80 nM) of each HDL subpopulation and lipid-free APOA1 (10 μg/mL). PC export and CEC were quantified with or without induction of ABCA1 expression with mifepristone. Radiolabeled outer leaflet phospholipids (PLs) were exchanged with unlabeled POPC and sphingomyelin with MαCD. PC efflux was calculated as the percentage of radiolabel in the medium of the cells divided by the total radioactivity of the medium and cells. PC export and CEC were quantified as described in Methods in cells with and without induced expression of ABCA1. Results are representative of triplicate analyses. The p-values were calculated by two-way ANOVA with Tukey’s post-hoc tests.

To compare inner leaflet PL with outer leaflet PL as substrates for ABCA1, we used MαCD to selectively exchange radiolabled PLs in the cells’ membrane outer leaflet with unlabeled POPC and sphingomyelin from the cells’ medium (**Figure 2A**).^39^ Lipid exchange markedly reduced (>90%) the export of radiolabeled PC into the medium by both APOA1 and isolated HDLs. These observations strongly suggest that outer leaflet PLs are the major PL substrate for export by ABCA1.

In contrast to PC export, efflux of radiolabeled cholesterol to APOA1 or isolated HDLs that was mediated by ABCA1 was little affected by lipid exchange with MαCD (**Figure 2B**). Thus, lipid exchange did not nonspecifically perturb the function of ABCA1. As we previously demonstrated for small HDLs (a mixture of XS-HDL and S-HDL by calibrated IMA),^19^ small HDLs were the most potent promoters of cholesterol efflux by BHK cells that expressed ABCA1. Taken together, these observations support the proposal that small HDLs promote cholesterol export by the ABCA1 pathway by driving PLs from the outer leaflet of the plasma membrane into the extracellular milieu.

## Discussion

Our findings demonstrate that HDL-P is strongly and negatively associated with incident CAD risk in a large cohort of people with T1D. HDL-P was quantified by a method that precisely and accurately determines particle size and has been validated with gold nanoparticles and reconstituted HDL particles.^34^ Moreover, the relative abundances of the HDL subspecies quantified by IMA are in excellent agreement with those determined by orthogonal techniques.^17, 34^ The negative association of T-HDL-P with CAD remained strong after adjustment for multiple potential confounders, including HDL-C, APOA1, and other traditional lipid and clinical risk factors. CEC, whether measured as macrophage CEC or ABCA1 CEC, also strongly and negatively associated with CAD risk in multivariable models. However, CEC was not a significant predictor of risk in models adjusted for T-HDL-P. These observations support the proposal that T-HDL-P is more strongly linked to CAD risk than is CEC in T1D, likely because HDL particle concentration associates with cholesterol efflux by the ABCA1 pathway.

In our study, HDL-C and APOA1 related only weakly to T-HDL-P. Also, triglyceride levels, which strongly and negatively associate with HDL-C levels, correlated only weakly and negatively with T-HDL-P, as did albumin excretion rate. Furthermore, there was little or no correlation of T-HDL-P with white blood cell count (a measure of systemic inflammation) and other established CVD risk factors. These observations suggest that the factors that control the association of T-HDL-P with CVD are independent of HDL-C and other traditional lipid and clinical CVD risk factors.

Although the four sizes of HDL strongly and inversely associated with incident CAD risk in our study, the strongest predictor of risk was the concentration of the smallest HDL particles, XS-HDL-P, with a 75% reduction in CAD incidence per 1 μmol/L increase in concentration. The risk reductions observed for T-HDL-P and the other sizes of HDL were smaller, ranging from 20% to 28%. Importantly, the small HDLs (XS-HDL and S-HDL) constituted less than 30% of the total HDL particle concentration in participants with and without CAD, with XS-HDL making up only 5%-6% of the total HDL particle concentration. These observations support the idea that both concentration and size have major impacts on HDL’s cardioprotective functions. They also support the proposal that small HDLs are of central importance in promoting cholesterol efflux from macrophages by the ABCA1 pathway. Other proposed functions of HDL, such as its antioxidant potential, are also enriched in small, dense HDL particles^41^ and might play a role in cardioprotection, as might the properties of the different sizes of HDL. For example, larger HDLs appear to exert antiatherogenic effects on the endothelium.^42^ It is also possible that different HDL functions predominate in different clinical settings, depending on the underlying pathophysiology and the contributions of the different sizes of HDL to cardioprotection.

A key unresolved issue is whether PL from the inner or outer leaflet of the plasma membrane drives cholesterol efflux by ABCA1. Our observations strongly suggest that outer leaflet PLs are the major substrate (though this scenario contrasts with the widely accepted alternating access model^22^). PL export by ABCA1 into the extracellular milieu then drives cholesterol efflux, likely because of a concentration gradient between the high concentration of free cholesterol in the plasma membrane and the cholesterol-free phospholipid exported by ABCA1.^21^ Consistent with this model, isolated small HDLs were much more efficient than larger HDLs at promoting phospholipid export from the outer leaflet of the plasma membrane.

Our findings suggest a mechanism for the link between small HDLs and CVD prevention. According to our flipped ends model,^20^ small HDLs can interact with ABCA1 more effectively than larger HDL particles because the C-termini of their two APOA1 molecules are flipped off the particle surface. In contrast, the C-termini of APOA1 in larger HDLs are bound to lipid. The enhanced ability of ABCA1 to promote PL efflux in turns enhances cholesterol efflux. Because small HDL particles promote cholesterol efflux more effectively than larger HDL particles, they might be a better metric for assessing CVD risk than the current clinical metric, HDL-C, at least in some populations.

Translational studies have reached different conclusions about the contribution of HDL’s size to CVD risk, but this dissonance may reflect the methods used to quantify HDL. As in our study, the Women’s Health Study, which used NMR to quantify HDL, found that low levels of small and very-small HDL particles associated with higher CVD risk.^43^ T-HDL-P quantified by NMR was also a better predictor of residual CVD risk in statin-treated people than was HDL-C in the JUPITER study.^16^ However, NMR measurements of HDL-P disagree with the size distribution of HDLs determined by 2-D gel electrophoresis, size-exclusion chromatography, and calibrated IMA.^44^ Moreover, two different NMR methods yield a stoichiometry of APOA1 per HDL particle of ∼1.5 and ∼7, respectively.^34, 45^ Both estimates are inconsistent with the currently accepted model for HDL’s structure, which posits 2-5 APOA1s per particle.^46^ Indeed, the correlation of T-HDL-P with plasma APOA1 measured by calibrated IMA (r=0.18, **Table 3**) is much lower than that of the Nightingale NMR method, which showed a strong correlation (r=0.87),^45^ suggesting that the NMR method quantifies APOA1 levels rather than HDL particle concentration.

Our observations may have therapeutic implications. The notion that HDL is not in the causal pathway for atherosclerosis was based largely on the relationship between CVD risk and plasma HDL-C levels in Mendelian inheritance studies^6, 47^ and the demonstration that two types of drugs that elevate HDL-C—niacin and cholesteryl ester transport protein inhibitors—failed to reduce CVD risk in statin-treated patients.^7, 8^ However, both drugs increase the size of HDL to a much greater degree than they raise the levels of APOA1 and/or T-HDL-P.^48–50^ Moreover, quantification of HDL by 2-D gel electrophoresis demonstrated that the concentrations of small HDL particles decreased in people treated with niacin or a CETP inhibitor ^50, 51^, suggesting that size and particle concentration are more important metrics of HDL’s cardioprotective effects than cholesterol content. Indeed, we estimated that extra-small HDL particles carried only ∼6% of the cholesterol in total HDL,^34^ which is consistent with the failure of HDL-C to associate with CVD risk in our study. CSL-112, a reconstituted HDL particle currently in a Phase III clinical trial for CVD event reduction in subjects with acute coronary syndrome^52^ increases small HDL and ABCA1-mediated CEC may provide a more promising strategy.^53^

Strengths of our study include the large, well characterized cohort of participants with T1D, the lengthy follow-up, the high rate of CAD events, the prospective assessment of CAD events, and the use of validated methods to quantify HDL-P and CEC. A limitation is that the participants we followed were not representative of the T1D population in the US because they were predominantly non-Hispanic white. However, the cohort reflects the demographics of the T1D population in Allegheny County, PA and the low incidence of T1D in racial minorities when the EDC study population was recruited. A key issue for future studies is whether small HDLs are strong predictors of risk in populations with type 2 diabetes and in non-diabetic populations.

In conclusion, we demonstrated that HDL-P levels strongly and negatively associate with incident CAD risk in people with T1D. The concentration of the smallest HDL particles− XS-HDL-P−was the strongest independent predictor of risk. Although CEC also strongly and negatively associated with CAD, it no longer predicted risk in models that included T-HDL-P, indicating that CEC was not independent of T-HDL-P. Our findings support the proposal that small HDLs are cardioprotective in T1D because they promote phospholipid and cholesterol efflux via ABCA1.

## Data Availability

All data produced in the present work are contained in the manuscript.

## Abbreviations

AER: albumin excretion rate
BMI: body mass index
MαCD: methyl-α-cyclodextrin
CAD: coronary artery disease
CEC: cholesterol efflux capacity
CVD: cardiovascular disease
EDC: Epidemiology of Diabetes Complications
eGFR: estimated glomerular filtration rate
HbA1: glycated hemoglobin
HbA1c: hemoglobin A1c
HDL-C: high density lipoprotein cholesterol
HDL-P: HDL particle concentration
HR: hazard ratio
IMA: ion mobility analysis
L-HDL-P: large HDL-P
M-HDL-P: medium HDL-P
NMR: nuclear magnetic resonance
PC: phosphatidylcholine
PL: phospholipid
RETRO-HDL-C: Re-Evaluating The Role of HDL Cholesterol in coronary artery disease
S-HDL-P: small HDL-P
T1D: type 1 diabetes
T-HDL-P: total HDL-P
WBC: white blood cell
XS-HDL-P: extra-small HDL-P

## Acknowledgements

We thank the study participants and staff for their contributions to the *Epidemiology of Diabetes Complications* (EDC) study and *Re-Evaluating the Role of HDL Cholesterol in coronary artery disease* (RETRO HDLc) study.

## Data availability

The authors confirm that the data supporting the findings of this study are available within the article and/or its supplementary materials

## Supplemental Material

**Supplemental Figure 1.**
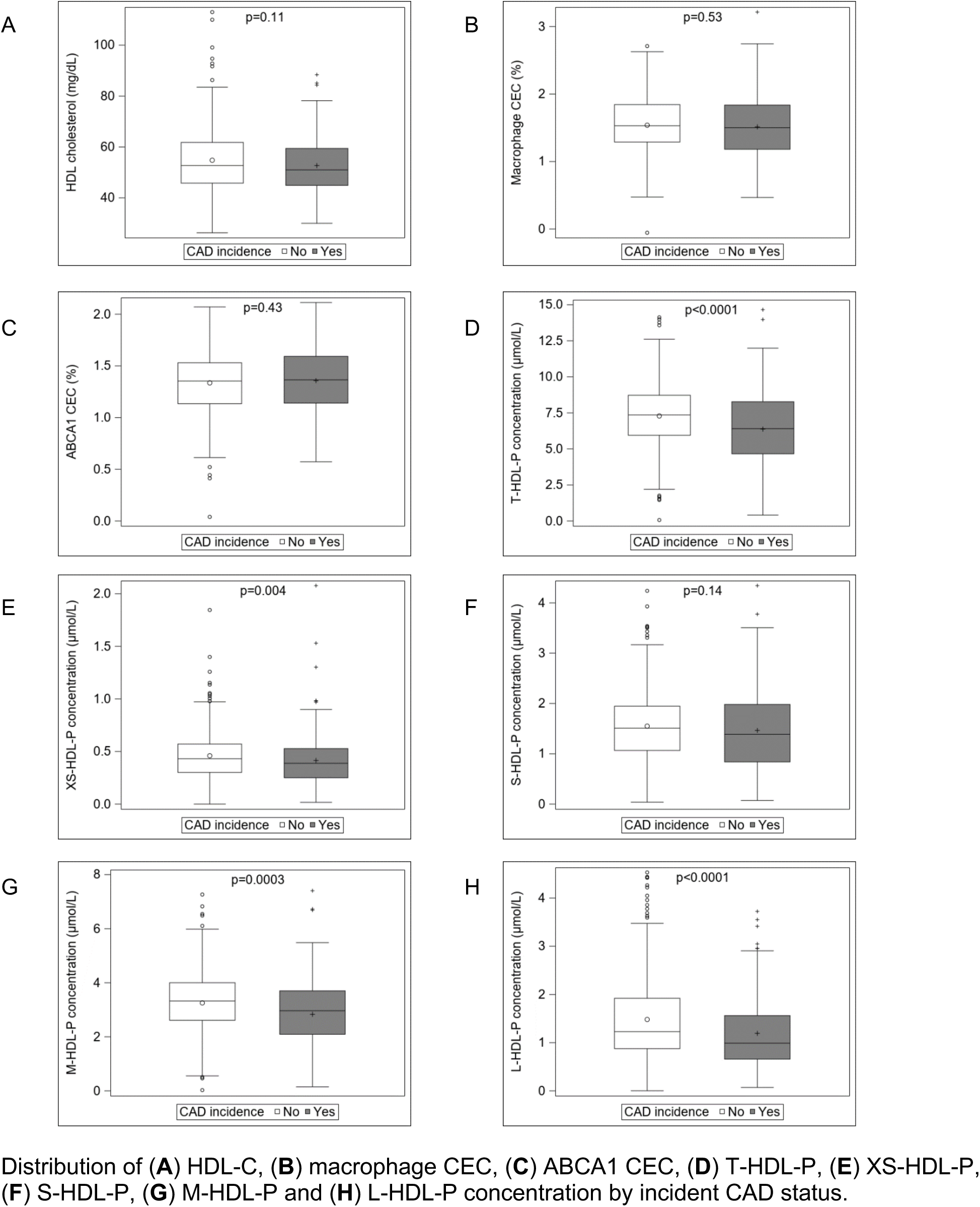
Distribution of HDL-C, serum HDL efflux capacity, and HDL-P concentration by incident CAD status

**Supplemental Figure 2.**
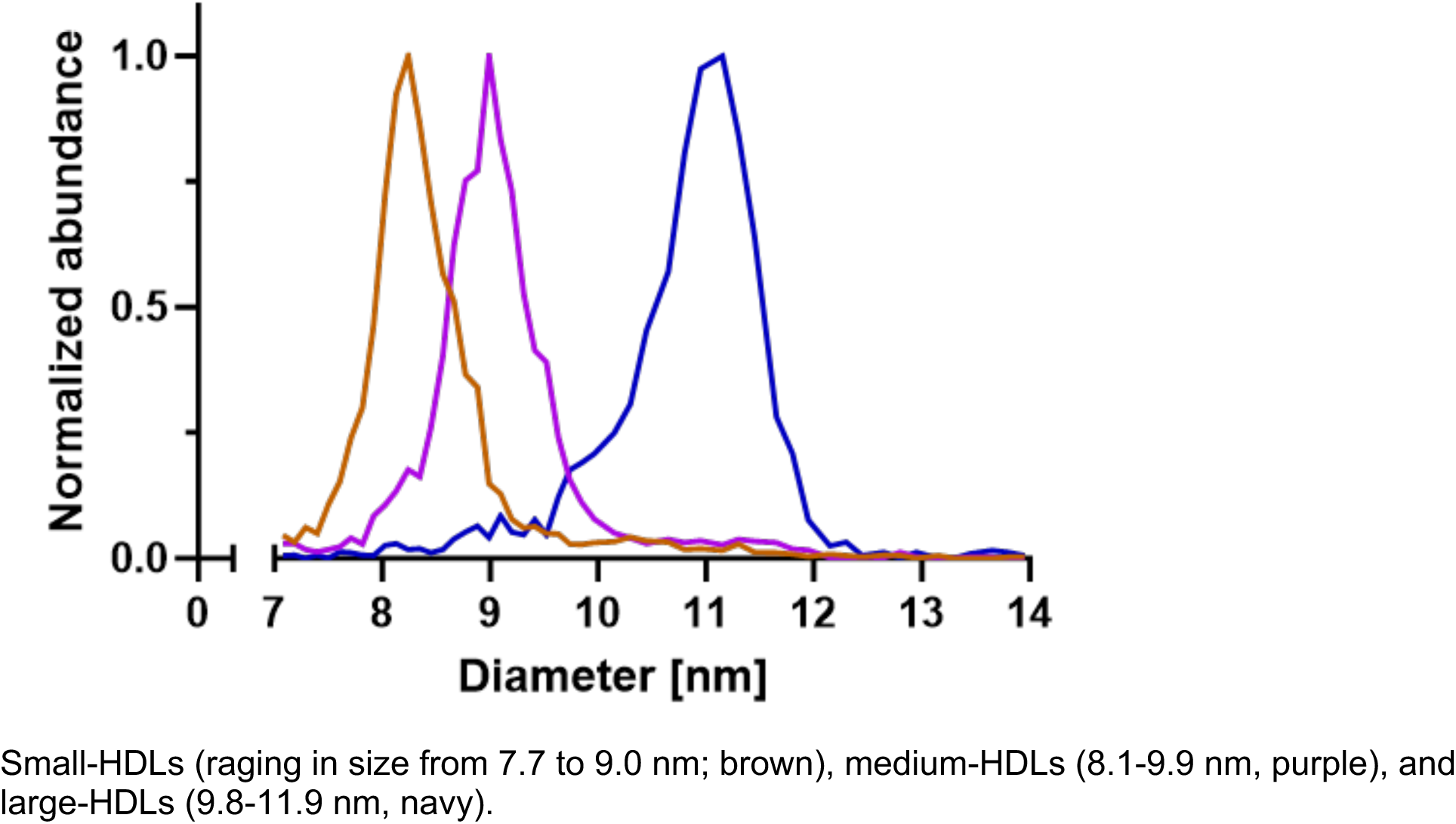
IMA profiles of small-HDL, medium-HDL and large-HDL isolated by gel filtration.

**Supplemental Table 1.** Cox proportional hazards models ^a^ (HR, 95% CI) for predicting incident CAD (n=502; 185 events) in models adjusted for LDL-C, triglycerides, and APOA1

| Main independent variable | Crude model | Model 1 | Model 2 | Model 3 |
| --- | --- | --- | --- | --- |
| HDL-C | **0.98 (0.97-0.99)** | 0.99 (0.98-1.01) | 0.99 (0.98-1.00) | 1.00 (0.98-1.01) |
| T-HDL-P | **0.85 (0.80-0.91)** | **0.89 (0.83-0.94)** | **0.88 (0.83-0.94)** | **0.88 (0.83-0.94)** |
| XS-HDL-P | **0.39 (0.19-0.81)** | **0.32 (0.16-0.63)** | **0.24 (0.12-0.48)** | **0.24 (0.12-0.48)** |
| S-HDL-P | 0.83 (0.68-1.03) | **0.79 (0.64-0.96)** | **0.80 (0.65-0.98)** | **0.80 (0.65-0.98)** |
| M-HDL-P | **0.77 (0.69-0.86)** | **0.82 (0.73-0.92)** | **0.80 (0.71-0.90)** | **0.80 (0.71-0.90)** |
| L-HDL-P | **0.61 (0.49-0.76)** | **0.78 (0.62-0.97)** | **0.79 (0.63-0.98)** | **0.79 (0.63-0.98)** |
| Macrophage CEC <sup>b</sup> | 0.97 (0.83-1.13) | **0.82 (0.71-0.95)** | 0.89 (0.77-1.04) | 1.02 (0.86-1.21) |
| ABCA1 CEC <sup>b</sup> | 1.06 (0.92-1.23) | 0.87 (0.75-1.01) | 0.85 (0.73-1.00) | 0.99 (0.83-1.18) |

<sup>a</sup> Model 1 allowed for HDL-C (for models where HDL-C was not the main independent variable), LDL-C, triglycerides and APOA1. Model 2 allowed for variables in Model 1 in addition to diabetes duration, sex, body mass index, smoking (time-dependent variable and fixed term), HbA1c, hypertension (time-dependent variable and fixed term), white blood cell count, estimated glomerular filtration rate, and albumin excretion rate.
Model 3 allowed for variables in Model 2 in addition to T-HDL-P (for models with HDL-C or CEC as the main independent variables) or macrophage CEC (for models with HDL-P as the main independent variables).
<sup>b</sup> HRs are per SD (macrophage CEC SD=0.44 and ABCA1 CEC SD=0.31)
Significant HRs and CIs in bold.

**Supplemental Table 2.** Correlation coefficients (p-values) between HDL metrics and clinical characteristics

|  | Cholesterol efflux capacity (%) |  | HDL-P concentration (μM/L) |  |  |  |  |
| --- | --- | --- | --- | --- | --- | --- | --- |
|  | Macrophage | ABCA1 | T-HDL-P | XS-HDL-P | S-HDL-P | M-HDL-P | L-HDL-P |
| Age | -0.09 (**0.03**) | -0.04 (0.38) | -0.09 (**0.04**) | -0.04 (0.31) | -0.13<br>(**0.003**) | -0.11<br>(**0.008**) | -0.05<br>(0.26) |
| Age at diabetes onset | 0.05 (0.20) | 0.008 (0.85) | -0.06 (0.14) | -0.03 (0.53) | -0.06 (0.16) | -0.10 (**0.02**) | -0.007<br>(0.87) |
| Diabetes duration | -0.13 (**0.003**) | -0.04 (0.40) | -0.06 (0.19) | -0.03 (0.54) | -0.09 (**0.04**) | -0.06 (0.17) | -0.06<br>(0.18) |
| Body mass index | -0.05 (0.21) | -0.009 (0.83) | -0.06 (0.14) | 0.01 (0.79) | 0.09 (**0.03**) | -0.002<br>(0.95) | -0.15<br>(**0.0005**) |
| Insulin dose per weight | 0.02 (0.57) | 0.03 (0.46) | -0.02 (0.57) | 0.07 (0.12) | 0.13<br>(**0.003**) | 0.03 (0.55) | -0.12<br>(**0.008**) |
| HbA<sub>1c</sub> | 0.22 (**<0.0001**) | 0.18 (**<0.0001**) | 0.04 (0.33) | 0.09 (**0.03**) | 0.13<br>(**0.002**) | 0.10 (**0.02**) | -0.08<br>(0.06) |
| Systolic blood pressure | 0.01 (0.79) | 0.06 (0.15) | -0.11 (**0.01**) | -0.04 (0.37) | -0.01 (0.78) | -0.09 (**0.03**) | 0.18<br>(**<0.0001**) |
| Diastolic blood pressure | 0.08 (0.05) | 0.10 (**0.01**) | -0.10 (**0.02**) | -0.02 (0.67) | 0.03 (0.43) | -0.04 (0.36) | -0.21<br>(**<0.0001**) |
| Pulse | 0.04 (0.39) | 0.07 (0.10) | -0.10 (**0.02**) | -0.06 (0.19) | -0.12<br>(**0.006**) | -0.08 (0.05) | -0.08<br>(0.08) |
| WBC | 0.004 (0.92) | 0.03 (0.47) | -0.15 (**0.0006**) | -0.05 (0.22) | -0.06 (0.16) | -0.13<br>(**0.002**) | -0.19<br>(**<0.0001**) |
| AER | 0.04 (0.37) | 0.07 (0.09) | -0.23 (**<0.0001**) | -0.06 (0.17) | -0.12<br>(**0.005**) | -0.23<br>(**<0.0001**) | -0.25<br>(**<0.0001**) |
| eGFR | 0.03 (0.48) | -0.03 (0.43) | 0.13 (**0.002**) | 0.07 (0.12) | 0.08 (0.06) | 0.13<br>(**0.002**) | 0.12<br>(**0.004**) |

AER, albumin excretion rate; eGFR, estimated glomerular filtration rate; WBC, white blood cell count.

